# Cohort profile: The Australian Children of the Digital Age (ACODA) longitudinal cohort study measuring the digital lives of Australians during early childhood

**DOI:** 10.64898/2026.05.09.26352795

**Authors:** Janelle E. MacKenzie, Daniel Johnson, Grace Sarra, Julian R. Matthews, Laura Martinez-Buelvas, Dana Trenaman, Steven J. Howard, Simon S. Smith, Susan Danby, Juliana Zabatiero

## Abstract

**Objectives:** The Australian Children of the Digital Age (ACODA) study is a longitudinal cohort study investigating the digital lives of Australians during early childhood. This paper presents a comprehensive description of the study protocol and overview of children’s digital technology use in the home at the first wave of data collection.

**Methods:** Caregivers of children aged 6-months to 5-years completed a survey that captured the availability and use of digital technology within the home, and child- and caregiver-related factors that may influence children’s digital technology use.

**Results:** A total of 3,388 caregivers from across all Australian states and territories completed the survey. Majority (98%) of children had digital technology and internet access within their homes. Most children (93%) used at least one device in the last year, with televisions, tablets, and mobile phones most frequently used (89%, 47%, 42%, respectively). Digital technology use started early, with 61% of children aged <1-year having used a television. A greater proportion of older children used devices, and for longer durations than younger children. Across all ages, daily time was longest on televisions (*M* = 1:20, *SD* = 1:14), tablets (*M* = 1:06, *SD* = 1:36), and mobile phones (*M* = 0:30, *SD* = 1:05). Digital technology was used most for entertainment and learning activities, and was used typically with a caregiver and in lounge/living rooms.

**Conclusions:** The ACODA study is the first longitudinal study to describe the digital technology use of Australians during early childhood and the context of this use. Data indicated that Australian children frequently used digital technology for entertainment and with their caregivers. Also, older children used digital technology more than younger children. Future waves allow for exploration of changes in children’s digital technology use over time, and associations with factors that may influence children’s digital technology use.

## Introduction

Australia is among the most digitally connected societies globally. National surveys report that 93% of Australian households have internet access at home and 83% of households have smart devices that connect to the internet [1]. By age 4, many children are regular users of tablets or smartphones for both learning and entertainment [2,3]. The COVID-19 pandemic further reinforced the centrality of digital technologies within the home, as families increasingly relied on devices for education, social connection, and leisure [4,5]. Recent reports indicate that television and video viewing, along with game playing, are the most common screen-based activities among children under 8 years of age [2]. Children use digital technologies for a variety of reasons, including entertainment, while parents complete tasks, as a reward for positive behaviour, and as a strategy to support emotion regulation when upset [2]. Taken together, this evidence highlights the central role of digital technologies in young children’s everyday lives. Nevertheless, digital disparities exist, with children in remote regions, Indigenous Australians, and lower-income households experiencing more limited access [6]. Those living in Australian capital cities experience greater digital inclusion than those in regional and remote locations, and a considerable digital gap persists between Indigenous Australians and non-Indigenous Australians, with lower access to digital technology being the critical issue for Indigenous Australians [7].

Within Australia, broad guidance is provided in the Australian 24-hour movement guidelines that recommend no sedentary screen use for children under 2 years, and no more than 1 hour per day for children between 2 and 5 years [8]. Similar recommendations are provided in other international guidelines [e.g., 9–11]. Indeed, there is evidence, mainly cross-sectional and focused on screen time, for associations between high screen use in early childhood and developmental risks, including delayed vocabulary [3], weaker language acquisition [12], disrupted sleep [13], and negative behavioural, cognitive and socio-emotional outcomes [14–16]. There is also evidence to suggest technology use is associated with improved learning capacity, productivity, and social competence, particularly when dimensions of use beyond duration, such as context and content, are considered [17–19]. Overall, the existing evidence does not yet provide clear guidance on children’s optimal use of digital technology.

The lack of clarity surrounding the benefits and risks of digital technology often leaves families confused and concerned, and experiencing anxiety and guilt, with little guidance on how to reconcile this uncertainty with the digital demands of the present and future. Current guidelines appear to be unrealistic for many families and do not account for the wide integration of technology within children’s home life. Recent years have seen a shift from a primary focus on screen time limits to a more nuanced consideration of context and content in advice to families. For example, the most recent policy statement from the American Academy of Pediatrics states that children’s media use cannot be considered through the lens of individual child behaviours or screen limits alone, and provides recommendations for families and professionals supporting them [20]. These are aligned with their previously launched ‘The 5Cs of Media Use’ framework, which helps parents foster healthy screen habits based on developmental needs [21].

Families require feasible, context-sensitive strategies that account for the diverse ways that technology is embedded in daily life [22,23]. However, we do not have a national snapshot of contemporary digital childhoods, comprehensively considering the contexts and conditions of young children’s digital engagement in the home. The Australian Children of the Digital Age (ACODA) longitudinal study provides the first nationally representative evidence of Australian children’s digital technology use in the home. ACODA follows over 3,300 families with children initially aged 6 months to 5 years, annually over 5 years. The study maps population-level patterns of children’s technology use, assesses children’s physical and mental health constructs as well as self-regulation abilities, and documents caregiver perspectives on both opportunities and challenges regarding children’s technology use. Analysis will be undertaken by Australia’s largest critical mass of interdisciplinary researchers in digital childhoods, bringing together expertise from multiple disciplinary backgrounds to inform understandings of children’s technology use. In generating rich longitudinal insights, ACODA delivers the evidence required to shape more equitable, realistic, and developmentally informed guidance in Australia and beyond.

This paper provides a comprehensive description of the ACODA longitudinal study, and an overall profile of the participant cohort, including description of the household socio-demographic characteristics, child digital technology use (i.e., what devices, where in the home, with whom, for how long, and for what purpose), and child-related factors that may be related with children’s technology use (e.g., self-regulation, mental health, social connectedness, sleep, physical activity, and disability or diagnosed condition status). These findings will provide the first step towards a description of contemporary digital technology use by Australian children and associated measures needed for the development of much needed contextual guidance for families.

## Methods

### Study Design and Setting

The ACODA longitudinal study was established in 2023 for the purpose of collecting population data to better understand digital technology use and how it potentially influences children’s wellbeing and development. The ACODA study was a foundational element of the Australian Research Council (ARC) Centre of Excellence for the Digital Child (herein referred to as the Digital Child Centre), a multi-year funded research centre with researchers from a range of disciplines interested in children’s digital childhoods (ARC Grant # CE200100022).

The ACODA study survey development was led by the Digital Child Centre’s chief investigators and a working group of centre members. These contributions of knowledge were from transdisciplinary researchers that spanned research domains from Early Childhood Education, Human-Computer Interaction, Psychology, Media Studies, Health Sciences and related disciplines.

### Recruitment and Data collection

Participants were recruited through invitations to participate via Digital Child Centre partners, including early learning centres, and child-focused organisations willing to promote the study. People who were the main caregiver of a child (or children): (1) aged from 6 months to 5 years 11 months, and (2) who lived with them for half of the time or more, were recruited into the study and completed the survey. These two requirements served as screening criteria. Each participant received a $50 e-gift card to compensate for their time. Wallis Social Research, a panel research company, was engaged to host the survey, validate participant responses, manage survey queries, and distribute the e-gift cards.

The study was promoted through the Centre’s partner organisations via social media, posters and flyers in centres/offices and newsletters. The Digital Child Centre general communication channels (e.g., social media, website) advertised the study, and Centre members promoted the study through their professional networks.

Baseline data collection for ACODA (Wave 1) took place from October 2023 to April 2024. A flow chart indicating the validation process for participant inclusion in the final cohort is provided in Figure 1. Data were initially removed if they were classified as bot-generated (based an assessment of data and skip errors and responses completed in an unrealistically short time), or if they did not meet the screening criteria during follow up calls. Wallis Social Research group called all respondents to confirm: (1) they were the only person in the house that completed the survey and (2) had only completed the survey once. For respondents who could not be contacted following multiple attempts, a Multivariate Validity Analysis was conducted to assess the likelihood of the respondents being valid (i.e., provided by a human and not a bot) with comparisons made against the data from verified respondents. If a likelihood of <0.35 was found, respondents’ data were removed. There were 5,250 initial survey respondents, and the final cohort was 3,388. The median time for survey completion was 29.77 minutes.

**Fig 1.**
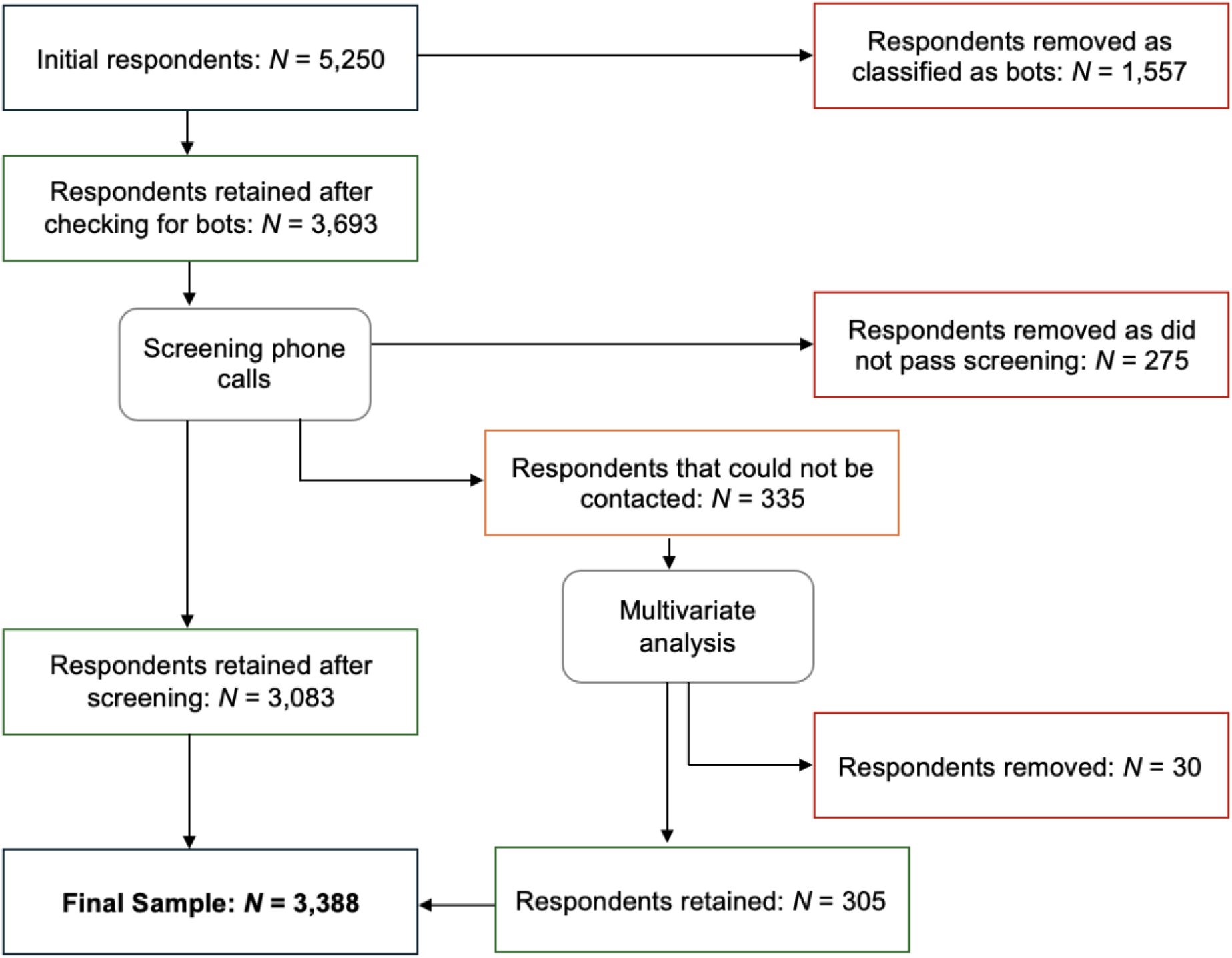
Flowchart of participant validation process for inclusion in the final cohort.

### Ethics Statement

The ACODA study was approved by Queensland University of Technology, Human Research Ethics Committee (#6427). Wallis Social Research hosted the ACODA survey, which included the approved participant information sheet and the digital consent process. All participants provided informed consent via a digital consent form prior to accessing the survey. Participants were advised that they could withdraw from the study at any time without prejudice or penalty. All data were securely stored on Wallis Social Research servers, institutional servers and anonymized before analysis.

### ACODA Survey Measures

The ACODA survey included six overarching sections (S1 Appendix). The length of the survey varied between participants as display logic was used to present appropriate measures based on the child’s age, as well as the presence of follow up questions if specific digital technology devices were present within the home and used by the child. Section 1 included screening items, basic caregiver information, and basic information regarding potential study child/ren to allow for inclusion and/or selection of the study child. Section 2 included items assessing the presence of digital technology in the house and the study child’s use of digital technology, assistive technologies, and internet access. Section 3 assessed caregiver perspectives on family and child’s digital technology use. Section 4 included items that assessed child-related variables that may influence digital technology use or health outcomes, such a child development, self-regulation, mental health, social connectedness, identity perception, physical activity, time outdoors, sleep, and play. Section 5 included items that assessed caregiver mental and physical health, and Section 6 included items that collected information regarding child and caregiver demographics and household context. In this profile paper we present results from a portion of the ACODA survey measures, with focus on describing the demographics of the ACODA cohort, ACODA children’s digital technology use, and child-related variables that may influence digital technology use. Data from Sections 3 and 5 of the survey are beyond the scope of this paper and not presented in the Results section.

#### Section 1: Screening information

Caregivers were asked to provide their date of birth (DOB), gender, number of adults and number of children that live in their home. For the children living in their home, the caregivers were asked to provide the DOBs, gender (i.e., male, female, non-binary, prefer not to say, or prefer to specify), and amount of time living in the home (i.e., 1 – Never, through 3 – About half the time, to 5 – All the time) for each child. If only one child fit the inclusion criteria for the study, they were selected as the study child. If there was more than one child that fit the inclusion criteria, the study child was randomly selected, and the caregiver was shown the DOB (of those they previously entered) of the selected study child. The caregiver was asked to indicate their relationship with the study child, and if the child lived elsewhere part of the time, the caregiver was asked to indicate with whom they lived.

#### Section 2: Digital technology

##### Digital technology availability and use at home (in the prior year)

Caregivers were asked to indicate their digital technology devices (from a list of 11 options: television, computer, mobile phone, tablet, e-reader, wearable device, handheld game, games console, virtual reality (VR) headset, internet-connected toy, and home assistant), and how many of each device, present in their home. Caregivers were asked to indicate if the study child used each device were present in the home in the prior year. For all devices the study child used in the prior year, the caregiver was asked to 1) indicate the room/s each device was used in, 2) indicate the people with whom the child used the device (response scale: 0 – no, 1 – yes; select all that apply, respectively), 3) whether the device could connect to the internet, and 4) if the child “had their own” device.

##### Child digital technology use (prior 7 days)

The caregivers who indicated that their child had used a device in the prior year were asked to indicate if their child used the device in the prior seven days. If they responded ‘yes,’ they were asked to 1) indicate the number of weekdays (i.e., 0 – 5 days) and weekend days (i.e., 0 – 2 days), and the average amount of time on these week- and weekend days (5 mins to 24 hrs) their child used the device.

For each device used by the child in the prior week, caregivers were asked to indicate the purposes of device use (i.e., learning activities, social connection, entertainment, playing games, creative activities, and/or rest or relaxation), and how often (i.e. from 1 – never, 2 – hardly ever, 3 - sometimes up to 6 – always) they used each device for that purpose. To capture “typical” purposes of use for each device, responses of 3 – sometimes to 6 - always were included in the descriptive analyses as it was deemed that these responses would portray “typical” purposes for using the device. Caregivers were not asked if their child used a television for creative activities, playing games, nor social connection, as it was deemed that parents typically do not perceive television to afford interactive creativity, play, nor social interaction. The items assessing children’s digital technology time use and purposes of use in the prior week were adapted from the Technology Use Questionnaire (TechU-Q), which has demonstrated good test-retest reliability, as well as face and construct validity [24].

##### Digital technology use outside the home

For all devices that the child did not use at home, the caregiver was asked to indicate (i.e., yes/no) if the child used those devices outside the home, such as at a family/friend’s house, school or public place.

##### Assistive technology

Caregivers were asked to indicate (i.e., yes/no) if their child used Assistive Technologies or Specialised Applications and provided a list of 10 technologies (ranging from vision support (braille displays)), to environment adaptations (lifts), and to assistive technology (mice or keyboards). Response options of ‘None,’ ‘Other (specify),’ ‘Don’t know,’ and ‘Prefer not to say’ were also offered.

##### Internet access

Caregivers were asked to indicate (i.e., yes/no) if they had internet access at home, and if so, if the study child ever accesses the internet at home. If internet access was not present in the home, the caregiver was asked to indicate if the study ever accessed free Wi-Fi when out of the home. If the study child accessed the free Wi-Fi when out of the home, the caregiver was asked to indicate the reasons for use and the location of use.

#### Section 3: Caregiver perspectives

##### Caregiver supervision

To assess caregiver supervision of their child’s digital technology use, the following question was posed: “In the past year, how often has anyone in your household supervised [study child] when using digital technology (either online or offline) in any of the following ways?” Responses included five supervision techniques (e.g., Sat beside them and helped them while they used digital technology) and an option for ‘Supervised in another way,’ and provided a rating scale of 1 – Never to 5 – Always. If the caregiver indicated that they ‘Supervised in another way’, they were presented with the opportunity to explain these other supervision practices with an open-ended response. Caregivers were asked if they had talked to their child about how to stay safe online in the past year (response scale: Yes, No, Don’t know, or Prefer not to say).

To assess caregiver monitoring and restriction of their child’s digital technology use, the following question was posed: “In the past year, have you or anyone in your household made use of any of the following…” with nine behaviours listed (e.g., Parental controls or other means of blocking or filtering some types of websites). The response options were Yes, No, or Don’t know. The caregivers were also asked (in the last year) how often they have “regulated the amount of time” and “supervised or controlled the ways” their child uses digital technology, and provided a response scale of 1 – Never to 5 – Always.

##### Information seeking

To assess caregiver information seeking, the following question was posed: “In the past year, which of the following sources have you looked for advice about how to help [study child] to stay safe when using digital technology?” and provided with a list of 10 sources, as well as options for ‘No, have not looked for or received any information or advice’ and ‘Other sources (specify).’ Caregivers were also asked to indicate the overall approach that they and their family had taken to the use of digital media and technologies in the past year. Four response options were provided - embrace, balanced approach, resistant to, and feel overwhelmed about – digital media and technologies.

##### Digital technology use and caregiver comfort

To assess specific digital technology use behaviours of the children and the caregiver’s comfort with their child’s use, 16 items (regarding specific behaviours, e.g., Playing educational games, making new friends online) were presented to the caregivers. The caregivers were asked to indicate if their child does any of the activities, and if so, the caregiver was prompted to indicate their comfort level with their child (at their current age) partaking in the behaviour on an 11-point scale (i.e., 0 – Not at all comfortable, to 10 – Completely comfortable).

#### Section 4: Child factors

##### Physical activity

One question was included to assess the study child’s physical activity, which captured their adherence to the 24-hour movement guidelines for all Australians [8] in the prior week. A description of the behaviours outlined in the guidelines for the child’s age were presented, and caregivers asked to report on how many days their child met the description.

##### Self-Regulation

Self-regulation items of the Child Self-Regulation and Behaviour Questionnaire [CSBQ; 25] were used to assess the behavioural (six items), cognitive (five items), and emotional (six items) applications of self-regulation, for children aged 3 years and above. The questionnaire poses the question “What is the child like?” and provides a list of statements for caregivers to indicate the accuracy of the statement for their child (i.e., from not true to very true). Average scores were calculated for each dimension of self-regulation. This measure has demonstrated good reliability, convergent validity with existing measures, and developmental sensitivity [25].

##### Mental Health

For children aged 1 to 4 years, Short-form (4-item) Patient-Reported Outcomes Measurement Information System (PROMIS) early childhood parent-report scales were used to measure the frequency of positive affect, anxiety and depressive symptoms [26] in the past week, using a 5-point Likert scale. These measures have demonstrated good content validity and developmental sensitivity [27].

For children aged 5 years, Short-form PROMIS parent proxy scales were used to assess positive affect [28; 4 items], anxiety (8-items) and depressive symptoms (6-items) [29] in the past week, using a 5-point Likert scale. Scores for all PROMIS mental health measures were calculated as the sum of the respective items.

##### Sleep

For children aged 6 months to 2 years, items from the Brief Infant Sleep Questionnaire [30] were included to assess sleep. Sleep constructs measured included number of nightly wakings (i.e., 0 to 10 or more), sleep onset difficulty (response scale: 5-point Likert scale from 1 – very easy to 5 – very difficult), duration of time study child usually takes to fall asleep (response scale: time windows ranging from 0-14 mins to 2 hours or more, and don’t know), duration of longest nighttime sleep period (response scale: hours, minutes, don’t know), duration of nighttime sleep (response scale: hours, minutes, don’t know), duration of awake time at night (response scale: time windows ranging from 0-14 mins to 2 hours or more, don’t know). These measures have demonstrated good concurrent validity and test-retest reliability [30].

For children aged 3 years or older, two short-form (4-item) PROMIS parent-report measures were included in the survey to assess sleep disturbance and sleep-related impairment [31] in the past week, using a 5-point Likert scale. Scores for these two measures were calculated as the sum of the four items. Validity as assessed by comparison to standard laboratory sleep study in a convenience sample demonstrated excellent precision [31].

##### Child development concerns

The caregiver’s concerns regarding their child’s development in five areas (i.e., fine motor, gross motor, communication/language, problem-solving, and social/emotional development) were assessed. Descriptors for developmentally appropriate behaviours were provided [drawn from the Ages & Stages Questionnaires, 32]. The descriptors were specific to age categories (i.e., 6-12 months, 1-2 years, 3-4 years, and 5 years) and caregivers were only shown descriptors that matched their child’s age. In future waves of data collection, these survey items will be replaced with the Child Regulatory Media Use scale [33] for study children aged 7 years and older. The caregivers were asked to rate their concern about their child’s development in the past year on an 11-point rating scale (i.e., 0 – *N*ot at all concerned to 10 – Extremely concerned), for each of the five development areas.

##### Social connectedness

To assess the study child’s level of social connectedness to those around them, the caregiver was asked to rate their child’s level of social connectedness to (1) family (e.g., caregiver, caregiver’s partner), (2) extended family (e.g., grandparents), and (3) friends on a 11-point rating scale (i.e., 0 – Not at all connected to 10 – Very connected).

##### Identity perception

The caregivers were asked to indicate whether they think (1) their child is considerate of other people’s differences, and (2) the media and other digital platforms portray a positive sense of their child’s identity (response scale: Yes, No, Don’t know, or Prefer not to say) [adapted from 34].

##### Time outdoors

To assess the amount of time the study child spent outdoors in the prior 7 days, the caregivers were asked to indicate (separately for weekdays and weekend days) the number of hours of the child’s free time was spent outdoors.

##### Play

The caregivers were asked to indicate to what extent they think their child learns through play when using digital technology, with a 5-point response scale from 1 – Never to 5 – Always.

#### Section 5: Caregiver factors

To assess caregivers’ mental and physical health within the prior 7 days, caregivers were asked to rate their (1) mental health (including your mood and ability to think), (2) satisfaction with social activities and relationships, and (3) physical health, on a 5-point rating scale from 1 – Excellent to 5 – Poor, with the additional option of ‘Prefer not to say’. Caregivers were also asked to rate to what extent they are able to carry out everyday physical activities (e.g., walking, climbing stairs, carrying groceries), on a 5-point rating scale from 1 – Completely to 5 – Not at all.

#### Section 6: Demographics & household context

##### Household context

Caregivers were asked to indicate the gender (i.e., male, female, non-binary, prefer to specify) of the other adults and children living in their home, as well as their relationship to the study child (e.g., father, mother, brother, sister, grandparent).

##### Demographics

Caregivers were asked to indicate their country of birth, the study child’s country of birth, and if the study child was born overseas, the year they arrived in Australia was requested. Caregivers were asked to indicate their ethnicity as well as the study child’s ethnicity with the list of options replicating the ABS collection of ethnicity and ancestry data [35]. In line with ABS data collection methods regarding ethnicity and ancestry, English was provided as the first ethnicity followed by other ethnicities in alphabetical order. Caregivers reported the primary language spoken at home and any other languages spoken, with the response options provided in line with ABS data collection methods [36]. Caregivers were asked to report if their child had a disability or diagnosed condition that requires support, and were provided a list of 12 disabilities or disorders (full list provided in S1 Appendix) as well as response options of no, other (specify), don’t know, and prefer not to say (response scale: select all that apply).

Caregivers reported on whether the study child had received or attended regular childcare from people or organisations other than them or their partner (response scale: yes, no). If yes, they were asked to indicate if these were: (1) Early childhood education and care services, (2) Formal or paid care from someone’s house, and/or (3) Family or friends, as well as how many days a week (i.e., 1-7 days) and how many hours in an average week (i.e., 1-168 hours).

Caregivers were asked to provide their relationship status (response scale: 6 options including married, divorced, widowed, other and prefer not to say), and highest education level (response scale: 10 options including Year 12 or equivalent, bachelor’s degree, postgraduate degree, other and prefer not to say). They were also asked to provide their employment status (response scale: 10 options including full-time, part-time, home duties, parental leave, other and prefer not to say), and the total number of hours typically worked in a week (response scale: 10 time ranges from 1-9 hours to 70 hours or more, and prefer not to say). Further details of all survey items are provided in S1 Appendix.

### Data analysis

All statistical analyses were undertaken using IBM SPSS Statistics (Version 29). Descriptive analyses were used to present an overview of demographic information, digital technology access within the home, and child physical and mental health, sleep, and self-regulation variables. To allow visual comparisons of the digital technology device use and the purposes of device use, proportional data for each age group are presented in percentages due to the uneven sample sizes across the age groups. Similarly, to account for the varying number of children that used each device, proportional data based on the number of children that used each device are presented when exploring with whom and where within the home children used each device.

#### Time child spent using digital technology

Descriptive analyses were used to describe the time children spent using digital technology devices. The total time spent for *each device* (e.g., television, tablet) used in the prior week was calculated as:

- *Total [device] weekday time* = *number of weekdays* x *average duration each day*
- *Total [device] weekend time* = *number of weekend days* x *average duration each day*
- *Total [device] weekly time = Total weekday time + Total weekend time*

The total time spent *across all devices* was calculated as:

- *Total weekly time across all devices* = *Total [television] weekly time* + *Total [tablet] weekly time* + *Total [mobile phone] weekly time* + *Total weekly time* [for 11 options]

Kruskal-Wallis H tests were conducted to test differences across age groups in weekly device use. This approach accounted for uneven sample sizes in the age groups, uneven numbers of children that used each device, and the positive skew in device use times. For significant findings, *post hoc* pairwise comparisons were conducted using Dunn’s [37] procedure with a Bonferroni correction with significance accepted at the *p* < .0033 level (to account to for 6 groups and 15 pairwise comparisons).

To account for the positive skew in the time use data, Mann-Whitney U tests were conducted to determine if there were significant differences in weekly time use between boys and girls. Distributions of the time use durations for boys and girls were similar, as assessed by visual inspection.

### Patient and public involvement

No members of the public were involved in the design, conduct, reporting, or dissemination plans of the present study.

## Results

The Wave 1 ACODA cohort included data from caregivers and their children. Comparisons have been made to the most current population data available from the Australian Bureau of Statistics [38], where appropriate. ACODA cohort characteristics are displayed in Tables 1 and 2. A high proportion of the ACODA caregivers identified themselves as the child’s mother (90.5%), and as such 91.1% of the caregivers were women. Sixty-seven percent of these caregivers were between 30 and 39 years of age, 65.3% were married, and a further 20.9% were in a de facto relationship or living with a partner. Over half of the caregivers had tertiary education, 34.1% were engaged in part-time employment, 29.9% were employed full-time, 10.2% were on parental leave from paid employment, and 10.4% were engaged with home duties.

**Table 1.**
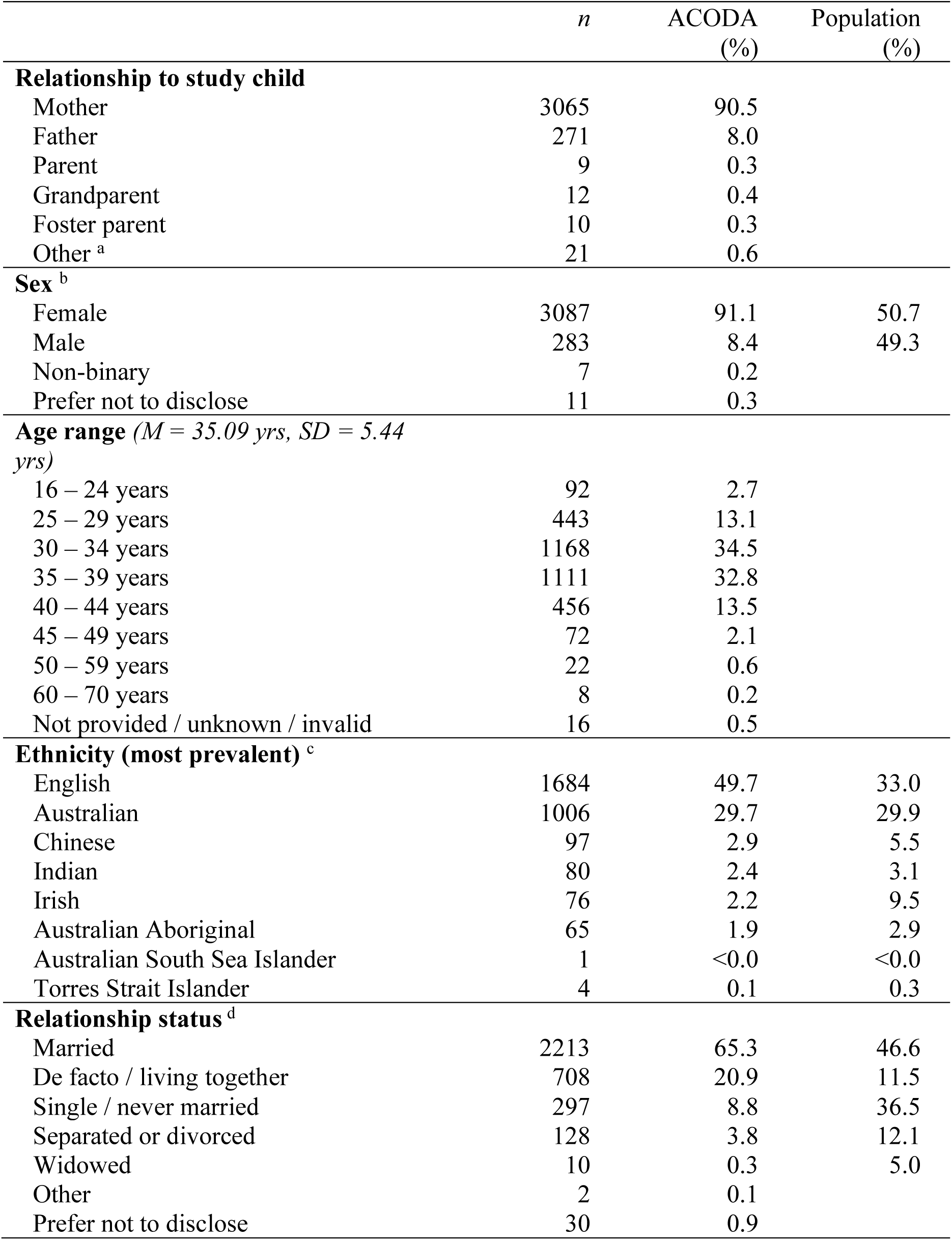
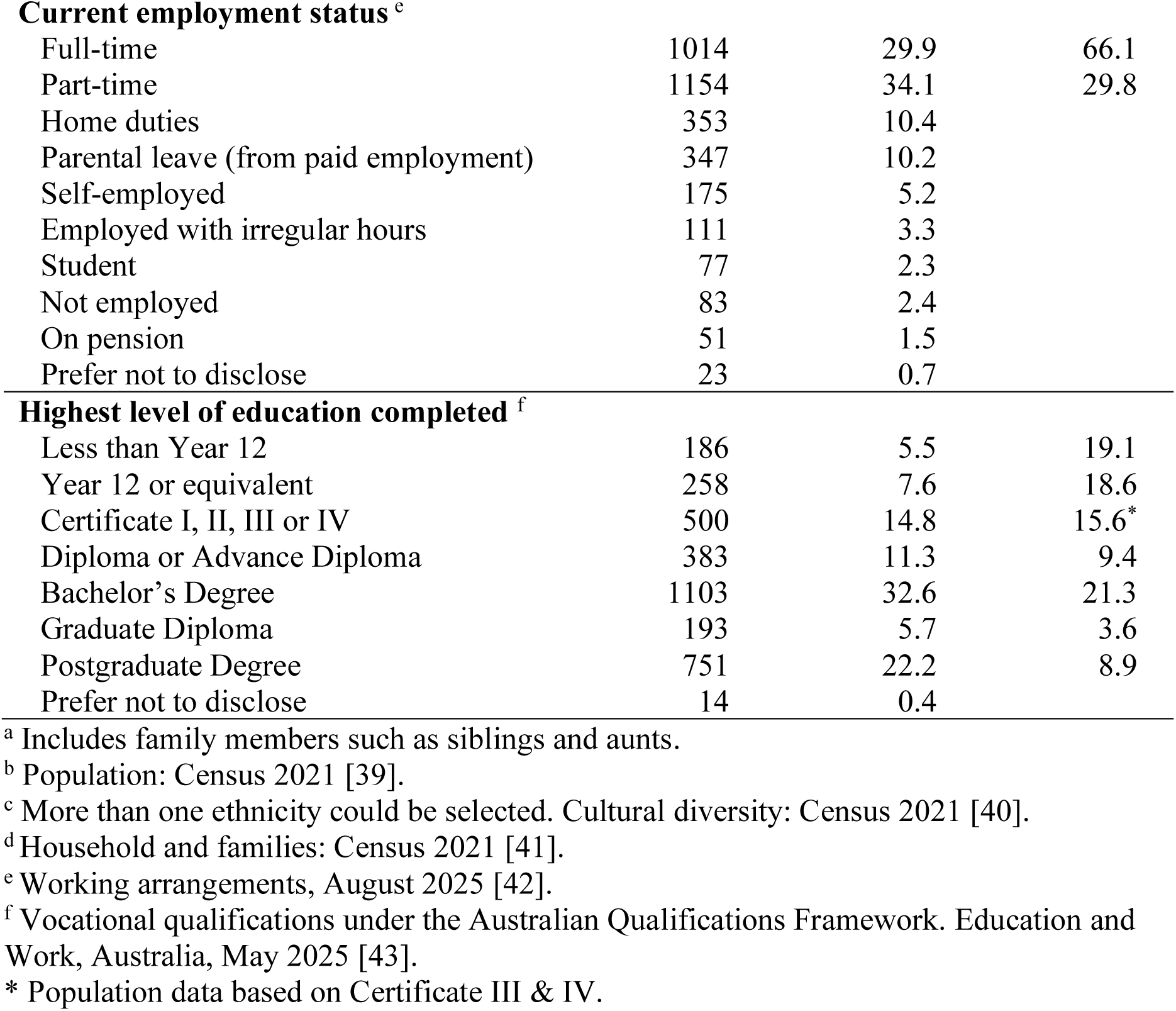
Wave 1 ACODA caregivers characteristics (*N*=3,388)

**Table 2.** Wave 1 ACODA children characteristics (*N* = 3,388)

|  | *n (%)* | ACODA<br>(%) | Population<br>(%) |
| --- | --- | --- | --- |
| **Sex <sup>a</sup>** |  |  |  |
| Male | 1738 | 51.3 | 51.0 |
| Female | 1648 | 48.6 | 49.0 |
| Non-binary | 2 | 0.1 |  |
| **Age Group** |  |  |  |
| 6 - 11 months | 321 | 9.5 |  |
| 1.0 - 1.4 years | 346 | 10.2 |  |
| 1.5 - 1.9 years | 364 | 10.7 |  |
| 2.0 - 2.4 years | 349 | 10.3 |  |
| 2.5 - 2.9 years | 382 | 11.3 |  |
| 3.0 - 3.4 years | 354 | 10.4 |  |
| 3.5 - 3.9 years | 329 | 9.7 |  |
| 4.0 - 4.4 years | 368 | 10.9 |  |
| 4.5 - 4.9 years | 275 | 8.1 |  |
| 5.0 - 5.4 years | 165 | 4.9 |  |
| 5.5 - 5.9 years | 135 | 4.0 |  |
| **Ethnicity (most prevalent) <sup>b</sup>** |  |  |  |
| English | 1561 | 46.1 | 33.0 |
| Australian | 1394 | 41.1 | 29.9 |
| Chinese | 79 | 2.3 | 5.5 |
| Indian | 79 | 2.3 | 3.1 |
| Italian | 73 | 2.2 | 4.4 |
| Australian Aboriginal | 120 | 3.5 | 2.9 |
| Australian South Sea Islander | 4 | 0.1 | <0.0 |
| Torres Strait Islander | 8 | 0.2 | 0.3 |
| **Disability or diagnosis <sup>c</sup>** |  |  |  |
| Present | 280 | 8.3 | 8.9 |
| Undergoing assessment | 16 | 0.5 |  |
| Suspected but undiagnosed | 7 | 0.2 |  |
| **Time living in caregiver's home** |  |  |  |
| All of the time | 3043 | 89.8 |  |
| Most of the time | 266 | 7.9 |  |
| About half the time | 79 | 2.3 |  |
| **Received childcare in the past year <sup>d</sup>** |  |  |  |
| Yes | 2748 | 81.1 | 59.0 |
| **Childcare received in the past year <sup>d</sup>** |  |  |  |
| ECEC services | 2607 | 76.9 | 43.3 |
| Family, friends, nanny / au pair | 938 | 27.7 | 31.6 |
| Formal or paid care at someone's home | 79 | 2.3 |  |

<sup>a</sup> Health of Children [44].
<sup>b</sup> More than one ethnicity could be selected. Population: Census 2021 [39].
<sup>c</sup> Child and Young People with Disability, 2022 [45].
<sup>d</sup> More than one childcare type could be selected. Australia's Children, Early Childhood Education and Care [46].

Just over half (51.3%) of the children were boys. The most prevalent ethnicities of the children were English and Australian. A disability or diagnosed condition was reported for 8.3% of the children. The majority of the children (89.8%) lived with the caregiver completing the survey all of the time. A high proportion (81.1%) of the children received childcare in the past year, with a high percentage of these (76.9%) attending an Early Childhood Education Centre (ECEC).

The characteristics of Wave 1 ACODA homes are presented in Table 3. Comparisons have been made to the most current population data available from the Australian Bureau of Statistics [ABS; 38], where appropriate. The ABS provides socio-economic indexes for areas within Australia (SEIFA) based on five yearly census data [47]. We utilised the Index of Relative Advantage and Disadvantage (IRSAD) that categorises Australian postcodes (geographical areas) into deciles, thus assigning a rating of 1 to 10 to each postcode (where 1 = most disadvantaged and 10 = most advantaged). The cohort lived in areas that spanned the range of IRSAD deciles, indicating a broad range of socio-economic advantage and disadvantage. However, half of the cohort resided in areas classified under deciles 7 to 10, indicating higher levels of socio-economic advantage.

**Table 3.** Wave 1 ACODA homes characteristics (*N* = 3,388)

|  | *n* | ACODA<br>(%) | Population<br>(%) |
| --- | --- | --- | --- |
| **State <sup>a</sup>** |  |  |  |
| New South Wales | 810 | 23.9 | 31.1 |
| Victoria | 596 | 17.6 | 25.6 |
| Queensland | 1254 | 37.0 | 20.5 |
| South Australia | 133 | 3.9 | 6.9 |
| Western Australia | 493 | 14.6 | 11.0 |
| Tasmania | 18 | 0.5 | 2.1 |
| Northern Territory | 24 | 0.7 | 1.0 |
| Australian Capital Territory | 60 | 1.8 | 1.8 |
| **Remoteness Classification <sup>b</sup>** |  |  |  |
| Major cities | 2558 | 75.5 | 73.0 |
| Inner regional areas | 533 | 15.7 | 18.0 |
| Outer regional areas | 257 | 7.6 | 8.0 |
| Remote areas | 25 | 0.7 | 1.1 |
| Very remote areas | 13 | 0.4 | 0.7 |
| Unknown | 2 | 0.1 |  |
| **Index of Relative Socio-Economic Advantage and Disadvantage <sup>c</sup>** |  |  |  |
| Deciles 1 - 2 | 359 | 10.6 |  |
| Deciles 3 - 4 | 510 | 15.5 |  |
| Deciles 5 - 6 | 773 | 22.9 |  |
| Deciles 7 - 8 | 857 | 25.3 |  |
| Deciles 9 - 10 | 875 | 25.8 |  |
| Unknown | 14 | 0.4 |  |
| **Number of adults in the home** |  |  |  |
| One | 318 | 9.4 |  |
| Two | 2777 | 82.0 |  |
| Three | 179 | 5.3 |  |
| Four or more | 114 | 3.3 |  |
| **Number of children in the home** |  |  |  |
| One | 1428 | 42.1 |  |
| Two | 1421 | 41.9 |  |
| Three | 394 | 11.6 |  |
| Four or more | 145 | 4.3 |  |
| **Main language spoken in the home** |  |  |  |
| English | 3078 | 90.9 |  |
| Mandarin | 27 | 0.8 |  |
| Spanish | 22 | 0.6 |  |
| Hindi | 18 | 0.5 |  |
| Aboriginal English | 12 | 0.4 |  |
| Punjabi | 12 | 0.4 |  |
| Other languages and prefer not to say | 219 | 6.4 |  |

<sup>a</sup> Estimated population as at September 2025 [48]
<sup>b</sup> Rural and remote health as at June 2023 [49]
<sup>c</sup> IRSAD - Based on postcode. Socio-Economic Indexes for Areas (SEIFA), Australia 2021[50]

The ACODA cohort resided across all Australian states and territories. The proportion of ACODA participants was similar to the expected proportion of the Australian population in the Australian Capital Territory and Northern Territory. A larger proportion of ACODA participants were from Queensland and Western Australia, and a lower proportion were from New South Wales, Victoria, South Australia, and Tasmania, when compared to census data (Australian Bureau of Statistics, 2024).

Three-quarters of the cohort (75.5%) lived in major cities of Australia, and a small proportion lived in remote areas (1.1%). There was a slightly greater proportion of ACODA families that resided in a major city, and slightly lower proportions residing in regional and remote areas, when compared to the proportions in the Australian population (Australian Institute of Health and Welfare, 2024). English was the main language spoken in 90.9% of the ACODA homes, with Mandarin, Spanish, and Hindi being the next most prevalent languages.

### Digital technology and internet access within the home

Table 4 describes the digital technology devices within ACODA homes. Nearly all homes had at least one television (97.3%) and mobile phone (96.6%). The next most prevalent devices within homes were computers (84.9%) and tablets (71.3%). Ninety-eight percent of homes had internet access, and 45.4% of the ACODA children accessed the internet in their home.

**Table 4.** Digital technology devices within ACODA homes (*N* = 3,388)

|  | *n* | *%* | *Median* | *Inter-quartile range* |
| --- | --- | --- | --- | --- |
| **Digital technology device <sup>a b</sup>** |  |  |  |  |
| Television | 3297 | 97.3 | 2 | 1 – 2 |
| Mobile phone | 3272 | 96.6 | 2 | 2 – 3 |
| Computer | 2877 | 84.9 | 2 | 1 – 2 |
| Tablet | 2417 | 71.3 | 1 | 1 – 2 |
| Wearable device | 1881 | 55.5 | 2 | 1 – 2 |
| Games console | 1398 | 41.3 | 1 | 1 – 2 |
| Home assistance device | 1069 | 31.6 | 1 | 1 – 2 |
| Handheld game device | 946 | 27.9 | 1 | 1 – 1 |
| e-reader | 670 | 19.8 | 1 | 1 – 1 |
| Internet connected toys | 345 | 10.2 | 1 | 1 – 2 |
| VR headset | 147 | 4.3 | 1 | 1 – 1 |
| Other digital device | 43 | 1.3 |  |  |
| No digital devices | 6 | 0.2 |  |  |
| **Internet access present** |  |  |  |  |
| Yes | 3319 | 98.0 |  |  |
| No | 58 | 1.7 |  |  |
| Don't know | 11 | 0.3 |  |  |
| **Study child accessed the internet** |  |  |  |  |
| Yes | 1537 | 45.4 |  |  |
| No | 1759 | 51.9 |  |  |
| Don't know | 23 | 0.7 |  |  |
| Did not respond | 69 | 2.0 |  |  |

<sup>a</sup> Present in the home in the prior year.
<sup>b</sup> All applicable devices could be selected.

Regarding ownership of digital technology, 20.3% of the children had their own tablet. However, very few children between 6-11 months and 1 year of age owned a tablet (i.e., 1.6% and 6.5% respectively). The proportion of children owning a tablet increased with each incremental year from 2 to 5 years (16.4%, 26.9%, 34.8% and 36.7%, respectively). Fewer ACODA children had their own internet connected toy (5%), television (4.3%), mobile phone (2.8%), handheld game device (1.7%), and games console (1.6%).

### Child digital technology use in the home

Ninety-three percent of the ACODA children used at least one digital technology device in the last year. The most used digital technology devices by ACODA children in the last year were televisions (88.9%), followed by tablets (47.4%) and mobile phones (41.6%). Of the 236 children that did not use a device (7% of the whole cohort), 152 of these children (64.4%) resided in IRSAD deciles 7 to 10 (i.e., postcodes with higher relative advantage), and a large proportion (84.3% of those that did not use a device) were 6-11 months or 1 year of age. However, as presented in Figure 2, 60.7% of 6-11 month olds in the whole sample used a television, and between 10-20% were using a mobile phone and a tablet.

**Fig 2.**
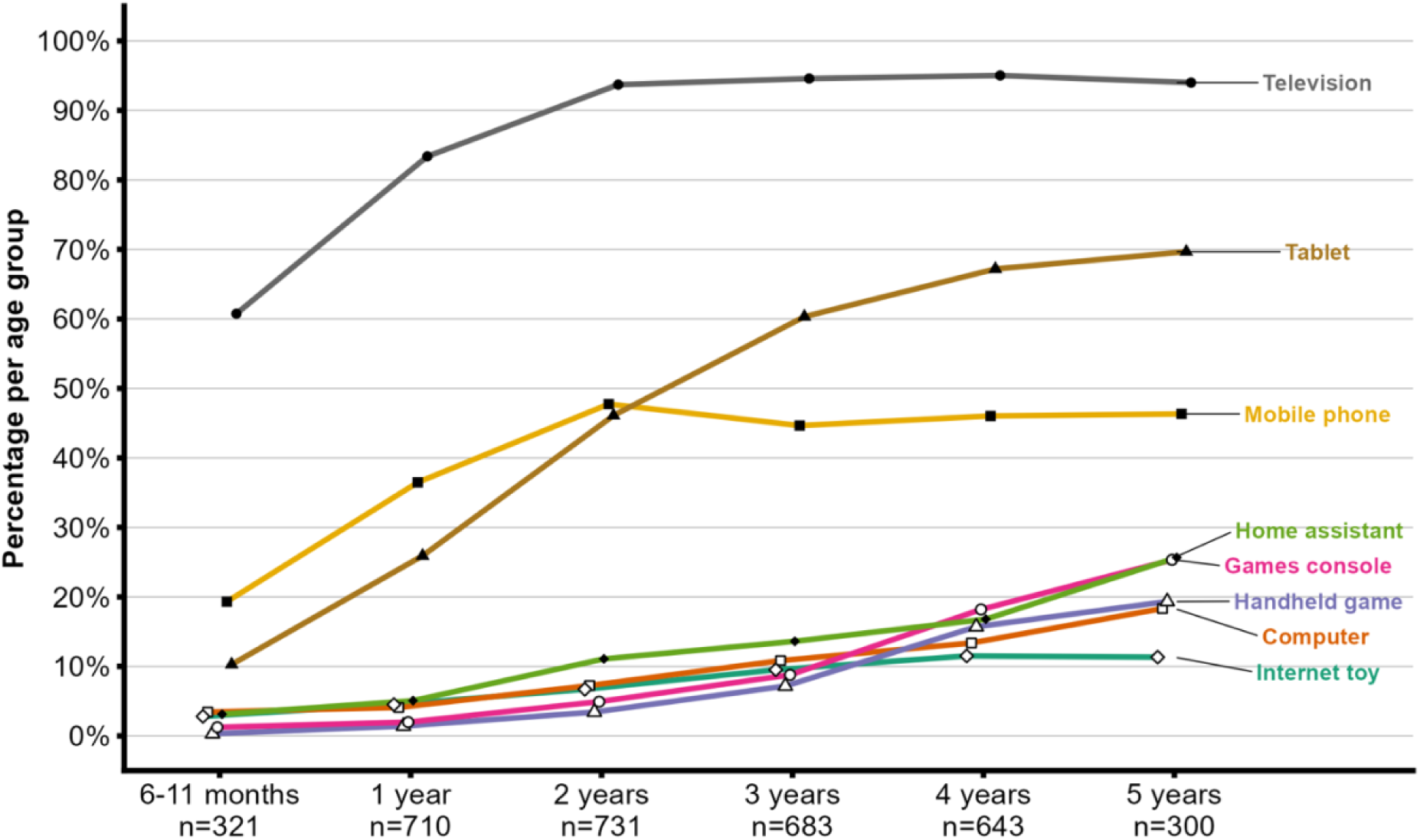
Digital technology device use by age group (*N* = 3,388)

As demonstrated in Figure 2, the proportion of children using devices was higher for children aged 2 years and older, when compared to those aged 6-11 months. The proportion of children using televisions and mobile phones remained consistent from 2 years to 5 years, whereas tablet use displayed a continued (albeit less steep) incline from 3 years onwards. There were gradual increases in the proportions of children using home assistant devices, games consoles, internet-connected toys and handheld games across the age groups. Wearable devices, e-readers and VR headsets were only used by a small proportion of the children across age groups (<4%).

### Time children spend using digital technology

The mean daily time for device use in the prior week *across all devices* was 1:56 (hours: minutes, *SD* = 2:16, *Range* = 0:00 – 33:43), however, there was a large amount of variability. Figure 3 presents the mean daily time by device type for all children that generally use each device (with those that did not use the device in the prior week recording values of 0 mins). The data were positively skewed, with a large proportion of children using devices for shorter durations. A small proportion of children used devices for longer durations, and it is likely that a proportion of children used multiple devices simultaneously (hence the maximum values exceeding 24 hours).

**Fig 3.**
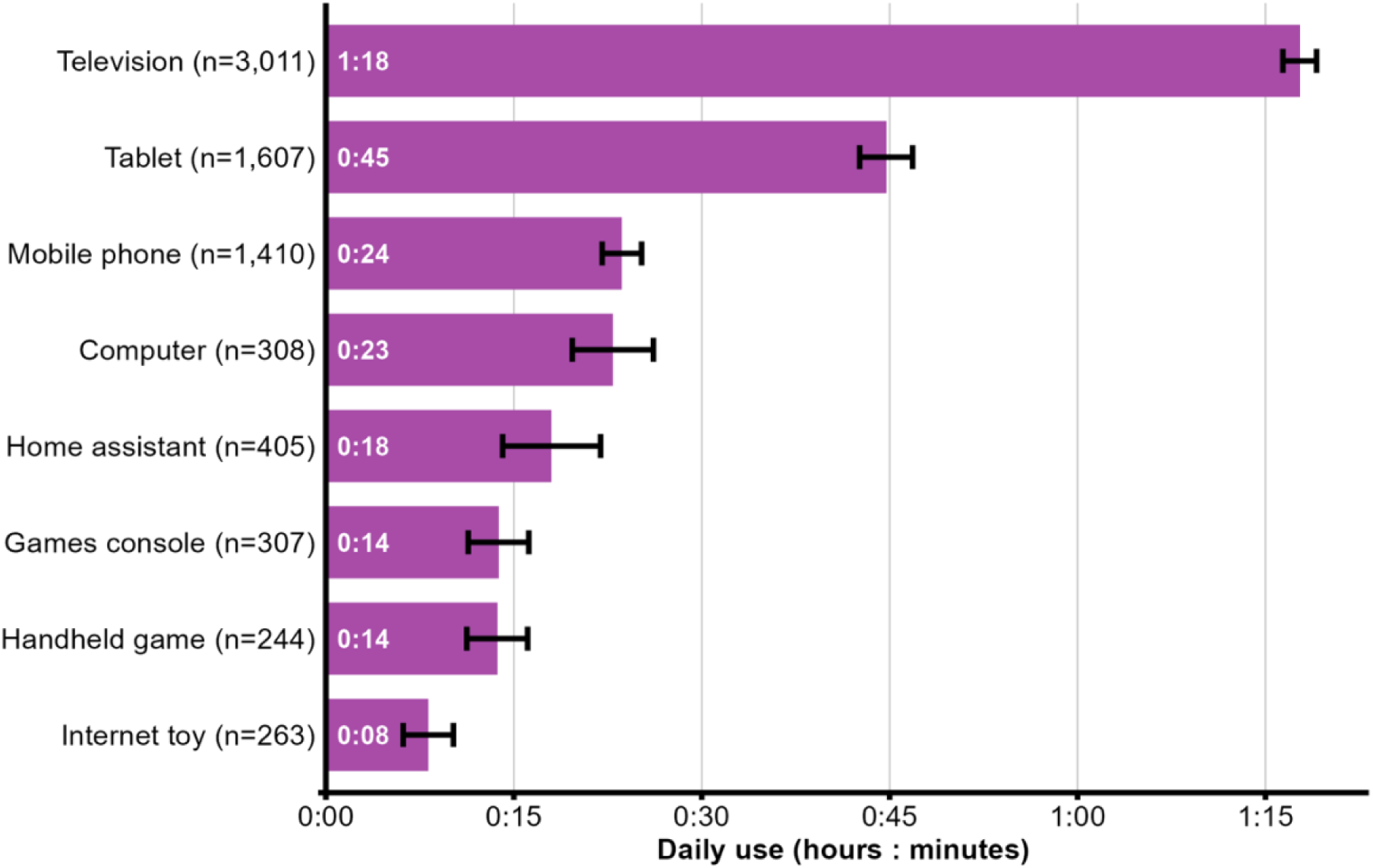
Mean daily time spent using digital technology devices (*N* = 3,388)

Ninety-one percent of the ACODA children used at least one digital technology device in the prior week, as 50 children that had used a digital technology device in the last year had not used one in the prior week. When considering the ACODA children’s digital technology device use in the past week, television was the most used device (86.2%), followed by tablets (34.0%), and mobile phones (33.0%). All remaining devices (i.e., home assistant, computer, games console, internet-connected toy, handheld game, wearable device, e-reader, and VR headset) were used by less than 10% of the cohort.

Table 5 displays the mean daily and mean weekly time for only those children that used the most common devices in the prior week. Televisions were used for the longest daily duration (*M* = 1:20, *SD* = 1:14), followed by tablets (*M* = 1:06, *SD* = 1:36), then mobile phones (*M* = 0:30, *SD* = 1:05). It is important to note the wide variability in these average device use times and positive skew of these data, due to the wide age range of ACODA children, indicating a large proportion with relatively short durations and a lower proportion with long durations.

**Table 5.** Time spent using top three devices by age group.

|  | *n* | % of age | Daily time |  | Weekly time |  |  |
| --- | --- | --- | --- | --- | --- | --- | --- |
|  |  |  | Mean | (SD) | Mean | (SD) | Median |
| **Television** |  |  |  |  |  |  |  |
| 6–11 months | 176 | 55 | 0:39 | (0:52) | 4:35 | (6:05) | 2:45 |
| 1 year | 570 | 80 | 1:00 | (1:14) | 7:02 | (8:36) | 4:00 |
| 2 years | 665 | 91 | 1:25 | (1:13) | 9:53 | (8:28) | 7:00 |
| 3 years | 632 | 93 | 1:27 | (1:13) | 10:07 | (8:30) | 7:00 |
| 4 years | 598 | 93 | 1:31 | (1:11) | 10:37 | (8:14) | 9:00 |
| 5 years | 276 | 92 | 1:39 | (1:19) | 11:31 | (9:16) | 9:00 |
| *All ages* | *2917* | *86* | *1:20* | *(1:14)* | *9:22* | *(8:36)* | *7:00* |
| **Tablet** |  |  |  |  |  |  |  |
| 6–11 months | 11 | 3 | 0:20 | (0:13) | 2:23 | (1:28) | 2:00 |
| 1 year | 102 | 14 | 0:46 | (1:38) | 5:23 | (11:28) | 1:53 |
| 2 years | 214 | 29 | 0:56 | (1:31) | 6:31 | (10:39) | 3:00 |
| 3 years | 280 | 41 | 1:05 | (1:35) | 7:32 | (11:03) | 4:00 |
| 4 years | 325 | 51 | 1:15 | (1:42) | 8:47 | (11:56) | 4:30 |
| 5 years | 136 | 45 | 1:19 | (1:28) | 9:16 | (10:14) | 4:45 |
| *All ages* | *1068* | *32* | *1:06* | *(1:36)* | *7:41* | *(11:11)* | *4:00* |
| **Mobile phone** |  |  |  |  |  |  |  |
| 6–11 months | 50 | 16 | 0:14 | (0:39) | 1:37 | (4:34) | 0:30 |
| 1 year | 219 | 31 | 0:14 | (0:24) | 1:35 | (2:47) | 0:35 |
| 2 years | 281 | 38 | 0:24 | (0:56) | 2:52 | (6:32) | 1:00 |
| 3 years | 216 | 32 | 0:33 | (1:07) | 3:53 | (7:47) | 1:30 |
| 4 years | 227 | 35 | 0:48 | (1:26) | 5:37 | (10:00) | 1:30 |
| 5 years | 110 | 37 | 0:42 | (1:27) | 4:57 | (10:06) | 2:00 |
| *All ages* | *1103* | *33* | *0:30* | *(1:05)* | *3:31* | *(7:37)* | *1:00* |

Three Kruskal-Wallis H tests were conducted to investigate whether there were significant differences across the age groups in weekly television, tablet, and mobile phone use. Overall, there were significant age trends in the proportion of the cohort and time spent using each of the top three device types. For those children using the top three devices, weekly time use was significantly higher for children in the older age groups (3+ years) than the younger age groups (≤1 year). This pattern was apparent for television, tablet and mobile phone use, albeit with slight differences across the age groups.

There was a statistically significant difference in weekly television time between the age groups, χ^2^(5, *N* = 2917) = 267.55, *p* <.001. Significant differences were found between the 6-11 month group (mean rank = 809.07) and all other age groups (*p* < .001), as well as significant differences between the 1 year group (mean rank = 1123.57) and all older age groups (mean ranks = 1531.21, 1585.99, 1636.42, 1717.00, consecutively) (*p* < .001). There were no significant differences between any other group combination (i.e., comparisons between 2, 3, 4, 5 years groups).

There was a statistically significant difference in weekly tablet time between the age groups, χ^2^(5, *N* = 1068) = 47.39, *p* <.001. The 1 year group (mean rank = 390.16) used tablets for significantly less time than the 3 years group (mean rank = 539.61) (*p* < .001), 4 years group (mean rank = 581.88) (*p* < .001), and 5 years group (mean rank = 610.58) (*p* < .001). A significance difference was also found between the 2 years group (mean rank = 485.13) and 5 years group (*p* = .003). There were no significant differences between any other group combination.

There was a statistically significant difference in weekly mobile phone time between the age groups, χ^2^(5, *N* = 1103) = 69.93, *p* <.001. Post hoc analysis revealed no statistically significant difference between the 6-11 month old group and the 1 year group, however both the 6-11 month old group (mean rank = 373.05) and the 1 year group (mean rank = 430.46) were significantly different from the 2 years group (mean rank = 556.25) (*p* = .003, *p* < .001, respectively), 3 years group (mean rank = 598.42) (*p* < .001), 4 years group (mean rank = 624.39) (*p* < .001), and 5 years group (mean rank = 623.93) (*p* < .001). There were no significant differences between any other group combination.

A Mann-Whitney U test was conducted to determine if there were differences in weekly time use across all devices between boys and girls and found statistically significant longer durations for boys (*Mdn* = 10:00) than girls (*Mdn* = 8:45), *U* = 1,134,314.00, *z* = -2.37, *p* = .018. A further three Mann-Whitney U tests were conducted to determine if there were differences in weekly time use between boys and girls for the top used devices. Weekly television time was statistically significantly higher for boys than girls, however, the median time for both genders was 7:00, *U* = 1,015,496.00, *z* = -1.97, *p* = .049. Weekly mobile phone time was statistically significantly higher for girls (*Mdn* = 1:25) than boys (*Mdn* = 1:00), *U* = 162,674.50, *z* = 2.06, *p* = .039. Weekly tablet time was not significantly different between boys (*Mdn* = 4:00) and girls (*Mdn* = 4:00), *U* = 143,561.50, *z* = 0.25, *p* = .804.

Of note, when inspecting use across all devices, slightly fewer children used devices on weekend days (*N* = 2865, 84.6%) compared to weekdays (*N* = 3025, 89.3%). However, the average daily time spent using devices on weekend days (*M* = 2:32) was higher than daily time on weekdays (*M* = 1:52).

### Purposes of device use in the prior week

To investigate the context within which the ACODA children were using digital technology within the home, the purposes for which the technology was used was explored. Overall, ACODA children used the top three devices to the greatest extent for entertainment, followed by learning activities. The purposes of use differed according to device use and age (see Figures 4 to 6).

**Fig 4.**
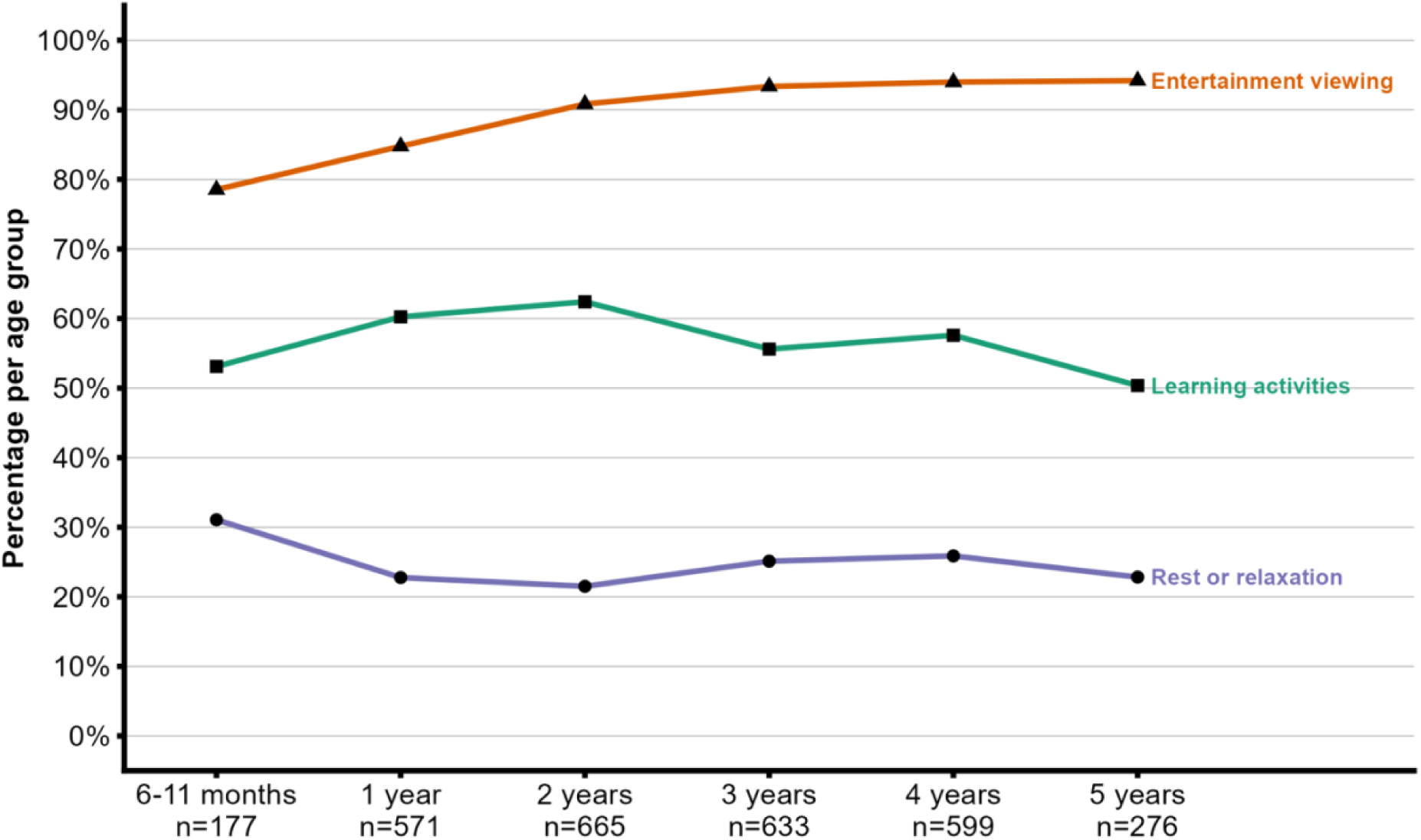
Purposes for using television by age groups (*N* = 2,921)

**Fig 5.**
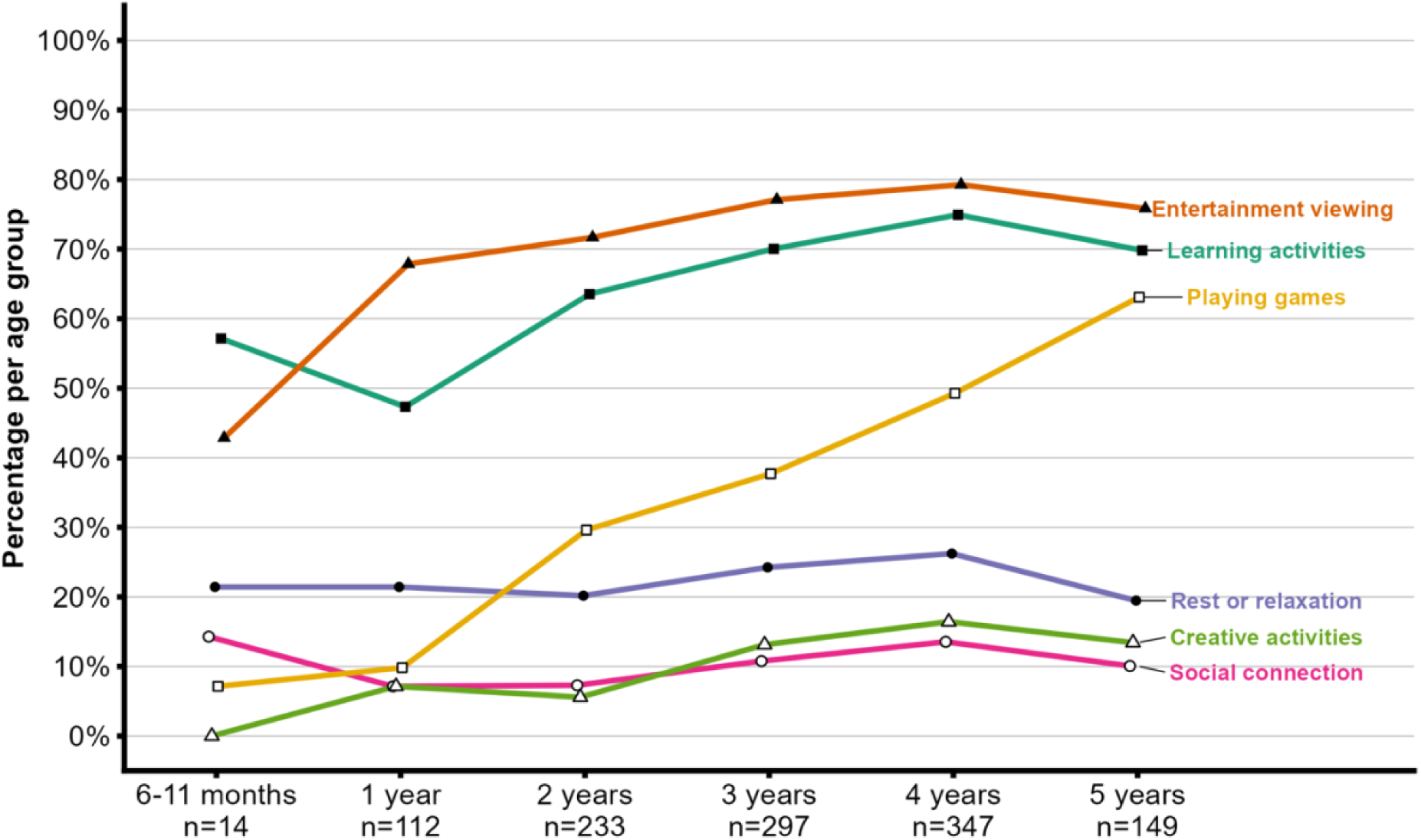
Purposes for using tablets by age groups (*N* = 1,152)

**Fig 6.**
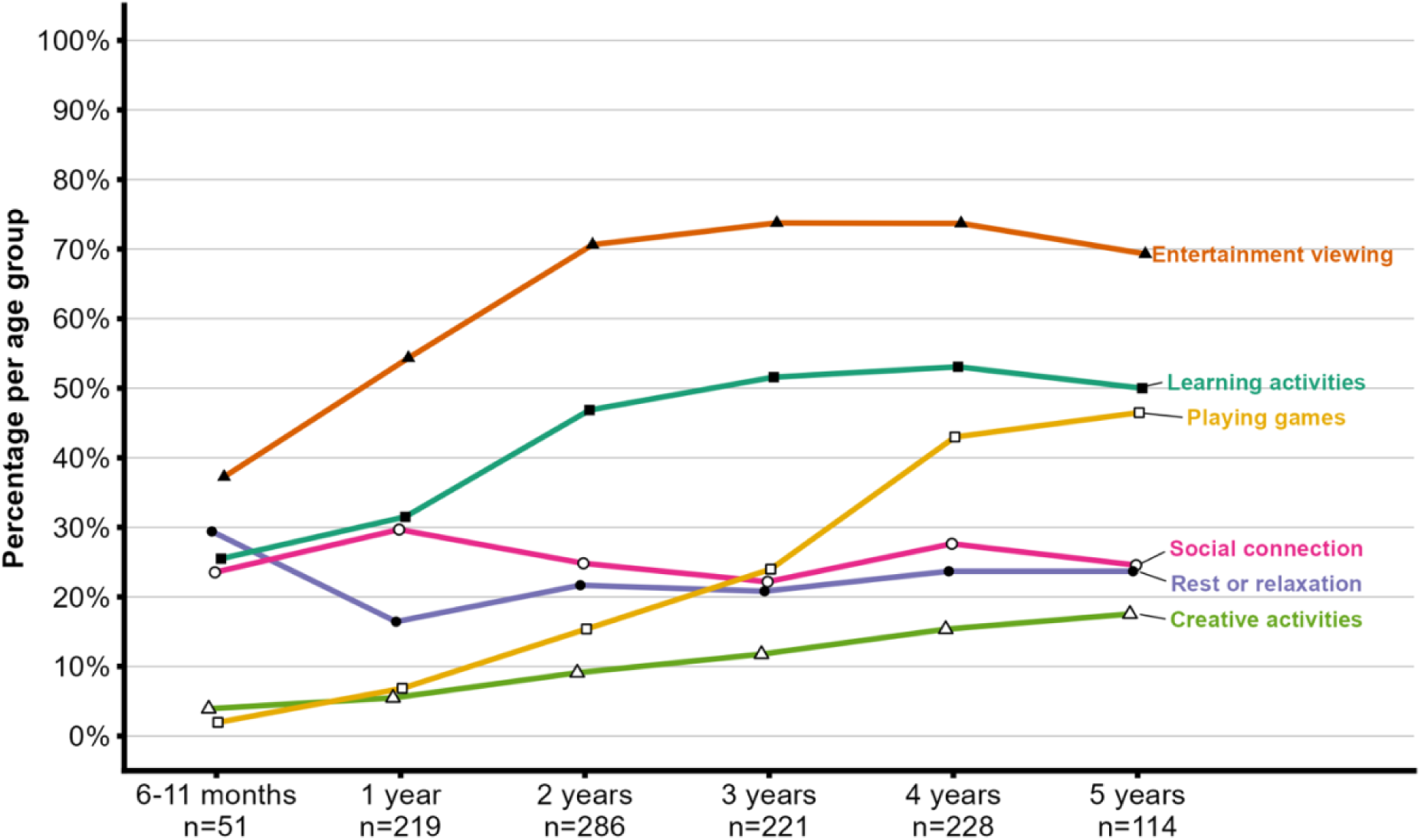
Purposes for using mobile phones by age groups (*N* = 1,119)

#### Television

Of the 2,921 children that used a television in the prior week, across age groups, the most common purposes were characterised as: (1) entertainment (over 78%); (2) learning activities (over 50%); followed by (3) rest or relaxation (over 20%) (as shown in Figure 4).

#### Tablet

Of the 1,152 children that used a tablet in the prior week, across age groups, the most common purposes were for: (1) entertainment, (2) learning activities, and (3) playing games. From 3 years onwards, over 75% of the children in each age group used a tablet for entertainment, with slightly lower proportions using a tablet for learning activities. The proportion of children playing games was more prevalent with older children (as shown in Figure 5).

#### Mobile Phone

Of the 1,119 children that used a mobile phone in the prior week, across age groups, the most common purposes were for: (1) entertainment, (2) learning activities, and (3) social connection. When considering children aged 2 years and older, over 70% used a mobile phone for entertainment, and over 45% for learning activities. Across all age groups, the proportion of children for each age group that used mobile phones for *social connection* was between 22% and 30%. A very small proportion of 6-11 months olds (2.0%) played games on a mobile phone, however, *playing games* was more prevalent with older children (as shown in Figure 6).

### Where and with whom children generally use digital technology

To further investigate the context within which the ACODA children use digital technology, where and with whom they use digital technology within the home (in the prior year) was explored. For all devices, caregivers reported that children mostly used them in the lounge room (Table 6). The top four rooms where a television was used were: (1) lounge room (97.3%); (2) other bedroom (12.1%); (3) playroom (8.0%); and (4) Child’s bedroom (5.0%). The high proportion of television use in the lounge room is in line with expectations given the stationary nature of televisions and common choice of placement. The top four rooms where a mobile phone was used were: (1) lounge room (74.6%); (2) other bedroom (21.3%); (3) kitchen (21.0%); and (4) child’s bedroom (20.6%). Similarly, the top five rooms where a tablet was used were: (1) lounge room (81.4%); (2) child’s bedroom (30.7%); (3) other bedroom (22.2%); (4) kitchen (21.8%); and (5) playroom (21.3%). The findings indicate that portable devices such as mobile phones and tablets were used in multiple rooms across the house, however, the lounge room remained the most prevalent location of use.

**Table 6.**
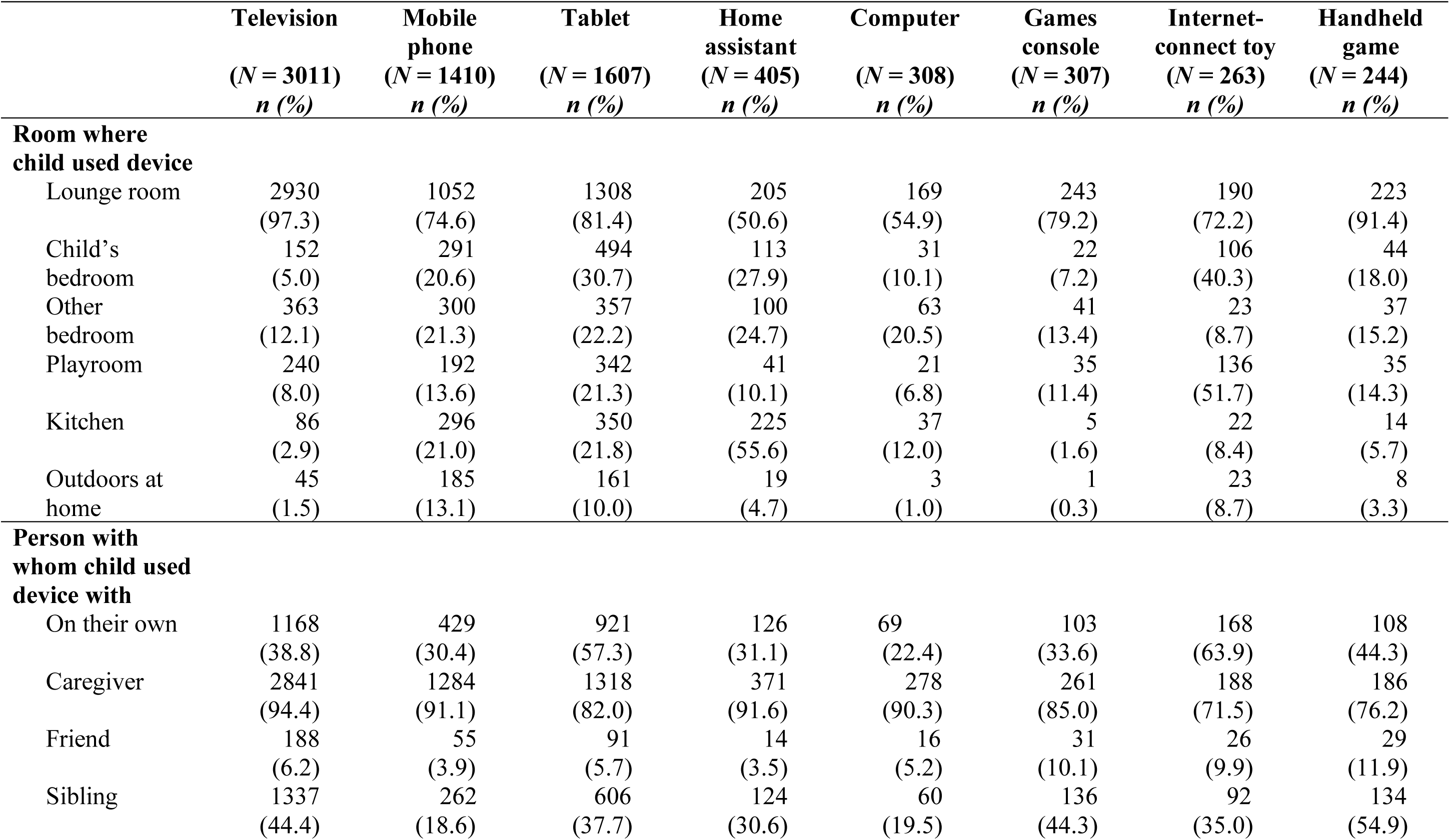
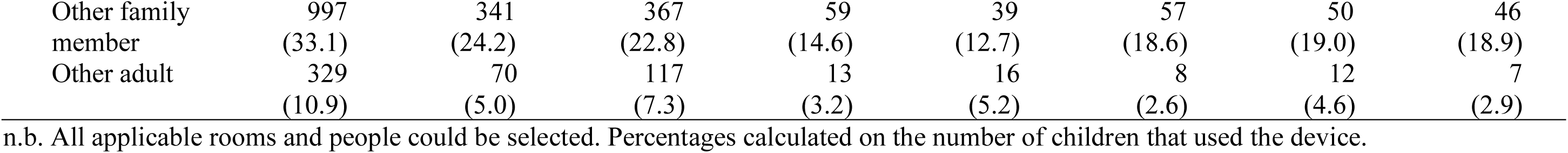
Where and with whom children generally use digital technology.

|  | **Television**<br>(*N* = 3011)<br>*n* (%) | **Mobile phone**<br>(*N* = 1410)<br>*n* (%) | **Tablet**<br>(*N* = 1607)<br>*n* (%) | **Home assistant**<br>(*N* = 405)<br>*n* (%) | **Computer**<br>(*N* = 308)<br>*n* (%) | **Games console**<br>(*N* = 307)<br>*n* (%) | **Internet-connect toy**<br>(*N* = 263)<br>*n* (%) | **Handheld game**<br>(*N* = 244)<br>*n* (%) |
| --- | --- | --- | --- | --- | --- | --- | --- | --- |
| **Room where child used device** |  |  |  |  |  |  |  |  |
| Lounge room | 2930<br>(97.3) | 1052<br>(74.6) | 1308<br>(81.4) | 205<br>(50.6) | 169<br>(54.9) | 243<br>(79.2) | 190<br>(72.2) | 223<br>(91.4) |
| Child's bedroom | 152<br>(5.0) | 291<br>(20.6) | 494<br>(30.7) | 113<br>(27.9) | 31<br>(10.1) | 22<br>(7.2) | 106<br>(40.3) | 44<br>(18.0) |
| Other bedroom | 363<br>(12.1) | 300<br>(21.3) | 357<br>(22.2) | 100<br>(24.7) | 63<br>(20.5) | 41<br>(13.4) | 23<br>(8.7) | 37<br>(15.2) |
| Playroom | 240<br>(8.0) | 192<br>(13.6) | 342<br>(21.3) | 41<br>(10.1) | 21<br>(6.8) | 35<br>(11.4) | 136<br>(51.7) | 35<br>(14.3) |
| Kitchen | 86<br>(2.9) | 296<br>(21.0) | 350<br>(21.8) | 225<br>(55.6) | 37<br>(12.0) | 5<br>(1.6) | 22<br>(8.4) | 14<br>(5.7) |
| Outdoors at home | 45<br>(1.5) | 185<br>(13.1) | 161<br>(10.0) | 19<br>(4.7) | 3<br>(1.0) | 1<br>(0.3) | 23<br>(8.7) | 8<br>(3.3) |
| **Person with whom child used device with** |  |  |  |  |  |  |  |  |
| On their own | 1168<br>(38.8) | 429<br>(30.4) | 921<br>(57.3) | 126<br>(31.1) | 69<br>(22.4) | 103<br>(33.6) | 168<br>(63.9) | 108<br>(44.3) |
| Caregiver | 2841<br>(94.4) | 1284<br>(91.1) | 1318<br>(82.0) | 371<br>(91.6) | 278<br>(90.3) | 261<br>(85.0) | 188<br>(71.5) | 186<br>(76.2) |
| Friend | 188<br>(6.2) | 55<br>(3.9) | 91<br>(5.7) | 14<br>(3.5) | 16<br>(5.2) | 31<br>(10.1) | 26<br>(9.9) | 29<br>(11.9) |
| Sibling | 1337<br>(44.4) | 262<br>(18.6) | 606<br>(37.7) | 124<br>(30.6) | 60<br>(19.5) | 136<br>(44.3) | 92<br>(35.0) | 134<br>(54.9) |
| Other family member | 997<br>(33.1) | 341<br>(24.2) | 367<br>(22.8) | 59<br>(14.6) | 39<br>(12.7) | 57<br>(18.6) | 50<br>(19.0) | 46<br>(18.9) |
| Other adult | 329<br>(10.9) | 70<br>(5.0) | 117<br>(7.3) | 13<br>(3.2) | 16<br>(5.2) | 8<br>(2.6) | 12<br>(4.6) | 7<br>(2.9) |

n.b. All applicable rooms and people could be selected. Percentages calculated on the number of children that used the device.

All devices were most prevalently used with a caregiver (Table 6). The people with whom the children most used a television with were: (1) a caregiver (94.4%); (2) a sibling (44.4%); and (3) on their own (38.8%). A mobile phone was used most with: (1) a caregiver (91.1%); (2) on their own (30.4%); and (3) another family member (24.2%). A tablet was used most with: (1) a caregiver (82.0%); (2) on their own (57.3%); and (3) a sibling (37.7%). There were low levels of device use with friends.

### Child factors

Table 7 presents mean scores for the physical activity and self-regulation variables, as well as norms for the self-regulation variables. Caregivers reported more than half (62.8%) of the ACODA children met the 24-hour movement guidelines on all seven days in the prior week (*M* = 6.95 days, *SD* = 1.67). The mean scores for ACODA children (aged 3 years and older) for behavioural self-regulation (*M* = 3.71, *SD* = 0.74) and cognitive self-regulation (*M* = 3.57, *SD* = 0.81) were slightly higher than the norms, whereas, the mean score for emotional self-regulation (*M* = 3.40, *SD* = 0.83) was slightly lower than the norms.

**Table 7.** Child factors.

| **Variable** | **Age Range**<br>**(yrs)** | **N** | **M (SD)** | **Range** | **Norms** | **M corresponding**<br>**percentile range** |
| --- | --- | --- | --- | --- | --- | --- |
| *Physical activity* |  |  |  |  |  |  |
| Days active per week | 0.5 – 5 | 3388 | 6.95 (1.67) | 0 – 7 |  |  |
| *Self-Regulation (CSBQ)<sup>a</sup>* |  |  |  |  |  |  |
| Behavioural Self-Regulation | 3 – 5 | 1625 | 3.71 (0.74) | 1 – 5 | 3.54 (0.90) |  |
| Cognitive Self-Regulation | 3 – 5 | 1625 | 3.57 (0.81) | 1 – 5 | 3.41 (0.84) |  |
| Emotional Self-Regulation | 3 – 5 | 1625 | 3.40 (0.83) | 1 – 5 | 3.48 (0.67) |  |
| *Sleep (PROMIS)<sup>b</sup>* |  |  |  |  |  |  |
| Sleep Disturbance | 3 – 5 | 1625 | 8.49 (3.28) | 4 – 20 |  | 56.9 – 59.1 |
| Sleep-related Impairment | 3 – 5 | 1625 | 9.44 (3.47) | 4 – 20 |  | 58.7 – 61.1 |
| *Mental Health (PROMIS)<sup>b</sup>* |  |  |  |  |  |  |
| Positive Affect (EC) | 1 – 4 | 2765 | 17.19 (2.12) | 8 – 20 |  | 46.6 – 50.0 |
| Anxiety (EC) | 1 – 4 | 2765 | 7.63 (2.53) | 4 – 20 |  | 54.9 – 57.5 |
| Depressive Symptoms (EC) | 1 – 4 | 2765 | 6.07 (2.19) | 4 – 20 |  | 54.5 – 57.6 |
| Positive Affect (PP) | 5 | 300 | 16.45 (2.24) | 10 – 20 |  | 48.3 – 52.0 |
| Anxiety (PP) | 5 | 300 | 15.72 (5.24) | 8 – 32 |  | 55.2 – 56.3 |
| Depressive Symptoms (PP) | 5 | 300 | 9.52 (3.15) | 6 – 24 |  | 53.2 – 54.9 |

<sup>a</sup> Preliminary Norms [25]
<sup>b</sup> Based on PROMIS T-score tables
n.b. EC = Early Childhood, PP = Parent Proxy

Table 7 presents the mean scores of the ACODA children for the PROMIS scales measuring sleep and mental health variables. The PROMIS T-score normative tables were consulted to identify the percentile range that the sample’s mean scores fell within (also reported in the Table 7). The mean scores for ACODA children (aged 3 years and older) for sleep disturbance (*M* = 8.49, *SD* = 3.28) and sleep-related impairment (*M* = 9.44, *SD* = 3.47) were within percentile ranges higher than the 50^th^ percentile, thus the ACODA children trended higher than the norms. The mean score for positive affect for ACODA children aged 1 to 4 years (*M* = 17.19, *SD* = 2.12) as well as those aged 5 years (*M* = 16.45, *SD* = 2.24) were close to the norm. However, the mean scores for anxiety and depressive symptoms were within percentile ranges higher than the 50^th^ percentile, thus the ACODA children trended higher than the norms.

## Strengths and Limitations

The study is the first to investigate young children’s (aged 6 months to 5 years) digital technology use in the home in a comprehensive and nuanced manner, moving beyond measures of time to consider the contexts in which digital technologies are used. Participants were drawn from all Australian states and territories, encompassing major cities through to remote locations, and representing a wide range of socio-economic circumstances. Embedded within a research centre dedicated to digital childhoods, the study was informed by multi-disciplinary expertise spanning multiple fields. The breadth of perspectives shaped both the study design and measurement approach. By capturing detailed information about children’s digital technology use alongside a broad range of factors influencing digital childhoods, the resulting dataset enables analysis and interpretation from diverse disciplinary perspectives.

Several limitations to the ACODA study should be noted. A relatively high proportion of participating caregivers had attained a tertiary education, which may limit the generalisability of the findings. While the predominantly online administration of the survey offered pragmatic advantages, it may have contributed to self-selection bias, particularly among caregivers with greater access to and familiarity with digital technologies. In addition, the study relied on caregiver reports of children’s digital technology use over the past week and year, as well as their psychological, social and behavioural functioning. As such, responses may have been influenced by recall or social desirability bias, with caregivers potentially reporting technology use in ways shaped by prevailing media narratives or social norms about “appropriate” use. Finally, as the survey was administered in English, families with lower levels of English proficiency may have been less likely to participate, introducing the possibility of further selection bias.

## Key Messages

- Digital technology devices and internet access were commonly available in ACODA homes.
- The majority of ACODA children used digital technology in the last year, and technology use started at an early age.
- Older children used digital technology devices more often and for longer durations than younger children. However, older children’s technology use became more variable indicating that some older children used digital technology for small durations whereas many others used digital technology for longer durations.
- Use of digital technology devices for entertainment was common and was higher by older children. Many children used digital technology for learning activities, and tablets and mobile phones were used by older children for playing games.
- Gender differences emerged across weekly time use variables, identifying that boys used televisions more than girls, whereas girls used mobile phones more than boys. However, weekly tablet use was not different between boys and girls.

## Conclusion

The ACODA study is the first longitudinal study to describe the digital lives of Australian children during early childhood and interrogate the context within which digital technology is used. Findings from the first wave of data collection indicate that children’s use of televisions, tablets, and mobile phones was prevalent. Use of these devices was relatively low for children under 1 year of age, with the proportion growing and then remaining consistent from 2 to 5 years for televisions and mobile phones, whereas the proportion of children using tablets gradually increased across all age groups. The time spent using these devices each week was significantly longer for older (3+ years) than younger children (≤1 year). Digital technology was frequently used with caregivers in lounge rooms, however, portable devices were used in various rooms within the home. Portable devices were often used for entertainment and learning activities from a young age, and increasingly used by older children for playing games. These patterns of use show the many reasons why children use devices and how this use may differ according to the child’s age. These findings emphasise the importance of increased investigation into the nuances of children’s digital technology use in the current ever-evolving digital era. The subsequent data collection waves will allow for reporting of longitudinal patterns of digital technology use as ACODA children grow older, and for further investigation of the associations between children’s digital technology use and other child-, caregiver- and home-related factors that may influence digital technology use.

## Data Availability

The authors advise that, due to ethical constraints, a de-identified data set cannot currently be made publicly available. Access requests may be directed to one of the ACODA study co-leads (Authors: DJ, GS, JZ).

## Acknowledgments

This research was supported by the Australian Research Council Centre of Excellence for the Digital Child through project number CE200100022. The authors would like to acknowledge the Australian Children of the Digital Age (ACODA) Longitudinal Study participants and their families for their time and involvement in the study. We would like to thank the ACODA Working Group members for their contributions to the development of the ACODA survey; The contributions of domain specific knowledge from across multiple disciplines created a comprehensive and transdisciplinary research project. We would like to thank the ACODA Project Manager, Claire Enkera, for her project support.

